# Assessing the recovery after cardiac surgery: Development and validation of the Fuwai-CRS (Fuwai-Cardiac Recovery Scale)

**DOI:** 10.64898/2026.03.03.26347484

**Authors:** Runchen Sun, Shen Lin, Zhongyu Jiao, Chenfei Rao, Xiaoting Su, Shuang Hu, Yan Zhao, Heng Zhang, Qiuling Shi, Sheng Liu, Wei Feng, Zhaoyun Cheng, Xiaoqi Wang, Chuzhi Zhou, Jue Wang, Yunpeng Ling, Zhenya Shen, Hai Tian, Zhe Zheng

**Affiliations:** National Clinical Research Center of Cardiovascular Diseases, Fuwai Hospital, National Center for Cardiovascular Diseases, Beijing, People’s Republic of China; State Key Laboratory of Cardiovascular Disease, Fuwai Hospital, National Center for Cardiovascular Diseases, Beijing, People’s Republic of China; Chinese Academy of Medical Sciences and Peking Union Medical College, Beijing, People’s Republic of China; Department of Cardiovascular Surgery, Fuwai Hospital, National Center for Cardiovascular Diseases, Beijing, People’s Republic of China; Key Laboratory of Coronary Heart Disease Risk Prediction and Precision Therapy, Chinese Academy of Medical Sciences and Peking Union Medical College, Beijing, People’s Republic of China; School of Public Health, Chongqing Medical University, Chongqing, People’s Republic of China; Department of Cardiovascular Surgery, Fuwai Central China Cardiovascular Hospital, Zhengzhou, People’s Republic of China; Department of Cardiovascular Surgery, Fuwai Yunnan Cardiovascular Hospital, Kunming, People’s Republic of China; Department of Surgical Intensive Care Unit, Fuwai Hospital Chinese Academy of Medical Sciences, Shenzhen, People’s Republic of China; Department of Cardiac Surgery, The First Affiliated Hospital of Wenzhou Medical University, Wenzhou, People’s Republic of China; Department of Cardiac Surgery, Peking University Third Hospital, Beijing, People’s Republic of China; Department of Cardiovascular Surgery of the First Affiliated Hospital & Institute for Cardiovascular Science, Soochow University, Suzhou, People’s Republic of China; Division of Cardiovascular Surgery, The 2nd Affiliated Hospital of Harbin Medical University, Harbin, People’s Republic of China; National Health Commission Key Laboratory of Cardiovascular Regenerative Medicine, Fuwai Central China Hospital Central China Branch of National Center for Cardiovascular Diseases Zhengzhou People’s Republic of China

**Author notes:** **Corresponding Author:** Zhe Zheng, MD, PhD, Department of Cardiovascular Surgery, Key Laboratory of Coronary Heart Disease Risk Prediction and Precision Therapy, National Clinical Research Center of Cardiovascular Diseases, State Key Laboratory of Cardiovascular Disease, Fuwai Hospital, National Center for Cardiovascular Diseases, Chinese Academy of Medical Sciences and Peking Union Medical College, No. 167 Beilishi Road, Xicheng District, Beijing 100037, People’s Republic of China. Contributed equally (Runchen Sun, Shen Lin).

**Keywords:** cardiac surgery, recovery, patient-reported outcome measure

## Abstract

**Background:** Cardiac surgery significantly improves clinical endpoints but imposes challenges in postoperative recovery. Assessing patient-reported outcome is crucial for optimal care. However, no cardiac surgery-specific tools currently exist to adequately capture postoperative recovery experience.

**Objectives:** To develop and validate a recovery scale after cardiac surgery (Fuwai-CRS).

**Methods:** This study was conducted from May 2023 to December 2024, comprising: (1) a qualitative study (Cohort 1) enrolling postoperative patients of cardiac surgery and medical staffs to develop the draft scale through literature review, semi-structured interview and Delphi consensus; and (2) a single-center prospective validation study (Cohort 2) to finalize the scale and evaluate psychometric properties.

**Results:** In Cohort 1, a 17-item draft Fuwai-CRS was generated based on literature review, semi-structured interview (40 patients and medical staffs) and a Delphi study (15 experts). In Cohort 2 (n=500), a 9-item Fuwai-CRS was finalized by data distribution assessment, hierarchical cluster and factor analysis, and its understandability, reliability, validity and responsiveness were found acceptable.

**Conclusions:** The Fuwai-CRS is a concise and valid tool for recovery assessment after cardiac surgery.

## Introduction

Improving patient recovery after cardiac surgery is a critical priority.^1^ While improved surgical techniques have substantially reduced procedural mortality over the past decades,^2,3^ the inherent physiological insult of median sternotomy, cardiopulmonary bypass, and myocardial manipulation continues to impose significant challenges in patient’s recovery experience, such as pain, sleep disorders.^4,5^ Paradoxically, this critical recovery phase remains underexplored, as traditional outcome metrics predominantly focus on mortality and major morbidity endpoints. Patient-reported outcome measures (PROMs) can provide complementary information by assessing the patient’s experience of symptom burden and functional status change over time,^6^ including generic tools in many types of surgery. Appropriate application of these tools have been shown to enhance recovery, improve daily functioning and quality-of-life,^7^ optimize healthcare utilization,^8^ and prolong survival.^9,10^

However, there is no widely accepted cardiac surgery-specific PROM. Previous studies used three categories of scales to assess recovery after cardiac surgery: (1) generic quality-of-life measures such as the 36-Item Short Form Survey (SF-36),^11^ and EuroQol five-dimensional questionnaire (EQ-5D)^12^; (2) generic surgery recovery scales such as the 15-item quality of recovery (QoR-15)^13^; and (3) cardiovascular-disease-specific instruments like the Kansas City Cardiomyopathy Questionnaire (KCCQ)^14^. Among these scales, only generic surgery recovery scale such as the QoR-15 fits the need of dynamic postoperative recovery assessment. However, QoR-15 was developed primarily in general surgery populations receiving general anesthesia, with limited representation of cardiac surgery patients^13^, which may not capture recovery challenges unique to cardiac surgery patients notably.^15^ In this prospective observational study, we aimed to: (1) develop a core-item, cardiac surgery-specific, postoperative recovery scale, named Fuwai Cardiac Recovery Scale (Fuwai-CRS); and (2) verify its reliability, validity and responsiveness.

## Methods

### Study design

This study comprised two prospective cohorts to develop and validate a cardiac surgery-specific recovery scale, Fuwai-CRS. We followed the processes of FDA guidance to develop and validate Fuwai-CRS, including scale item generation, cognitive debriefing and psychometric validation.^16,17^ The initial item pool was generated from literature review and qualitative interview with patients, cardiac doctors and nurses in the single-center Cohort 1 with the information saturation of sample size. The draft Fuwai-CRS was confirmed after consulting an expert panel (Delphi method).^18^ The final Fuwai-CRS was then determined through elimination of redundant items using three different methods (data distribution assessment, hierarchical cluster analysis and exploratory factor analysis) in the single-center Cohort 2. The understandability, reliability, validity and responsiveness were also validated in Cohort 2. The study was registered in clinicaltrials.gov (NCT05782036) and approved by the ethical review boards. All participants provided written informed consent.

### Participants

Cohort 1 contained patients who have undergone cardiac surgery, experienced clinical doctors and nurses in Fuwai Hospital. Patients underwent maximum variation sampling based on age, gender, disease type, education, and postoperative hospital stay.^19^ Clinical doctors and nurses have more than 5 years of experience in cardiac surgery. In Cohort 2, patients scheduled for elective cardiac surgery were screened. Exclusion criteria were: age under 18 years old, refuse to participate, prolonged postoperative mechanical ventilation exceeding 24 hours reoperation or perioperative death.

### Item generation

We generated a candidate item pool by inviting patients underwent cardiac surgery, cardiac doctors and nurses (Cohort 1) to qualitative interview **(Supplemental Methods)**. We also conducted extensive literature-based review from different countries for comprehensive and cross-cultural item generation.^20^ The experts evaluated each item’s clinical relevance using a 5-point Likert scale through two rounds of evaluation (Delphi method). Items with an average score of greater than or equal to the mean minus standard deviation across two rounds were retained in the provisional Fuwai-CRS.

### Cognitive debriefing and Item finalization

Patients scheduled for surgery in Cohort 2 completed the draft Fuwai-CRS at the following time points: the day before surgery, postoperative days 1-4 (D 1-4), and at week 2 (W2), week 3 (W3), 1 month (M1), and 3 months (M3) postoperatively. The timepoints were selected based on semi-structured interviews of Cohort 1 patients, in which we asked about their postoperative recovery changes over time in the follow-up period interview. They also completed the validated Chinese version of QoR-15.^21^ A subset of patients (n=21) was asked to complete cognitive debriefing to assess item comprehension, patient comfort, and acceptability after the draft Fuwai-CRS completion.^22^

Items were selected using three different methods to meet the required standard criteria, and redundant items were eliminated to generate the final Fuwai-CRS. First, we considered the floor and ceiling effects per item.^23,24^ The response distribution of each item was reviewed. We selected items that do not exceed 30% of ceiling or floor effects. Then we used hierarchical cluster analysis to identify groups of similar items.^25^ Eliminating the similar items decreased the length of the assessment and made it easier for patients to complete. Finally, exploratory factor analysis was conducted for item selection. Kaiser-Meyer-Olkin (KMO) test and Bartlett’s sphericity test values were examined to verify the application of exploratory factor analysis.^24^ A KMO value of 0.7 or higher and a p-value <0.001 from Bartlett’s sphericity test indicate the appropriateness of the data correlation analysis. The final items were chosen for factor selection by checking whether there were any items with a factor loading value >0.4 across several factors.^24^

### Reliability and validity

Cronbach’s α for internal consistency and the intraclass correlation coefficient (ICC) for test-retest reliability were evaluated for the final Fuwai-CRS. A value of 0.7 or higher for Cronbach’s α and ICC indicated good reliability.^24,26^ Criterion validity was determined by calculating Pearson’s correlation coefficients between Fuwai-CRS and QoR-15. We evaluated known-group validity by differentiating patients with varying statuses. Cohen’s d effect sizes and independent sample t-tests were used to compare scores across cardiac function [left ventricular ejection fraction (LVEF) ≥45 vs. LVEF < 45] and postoperative hospital stay (≥7 days vs. <7 days). We evaluated responsiveness by comparing baseline and postoperative scores using Cohen effect sizes and Standardized response mean.

### Statistical analysis

Continuous variables are presented as median [interquartile range (IQR)] and categorical variables are presented as proportions. Analyses were conducted on those with complete Fuwai-CRS data. All comparisons were two-sided, with statistical significance defined as P < 0.05. Analyses were calculated using R statistical software (version 4.0.3).

## Results

### Patient characteristics

We recruited 31 patients underwent cardiac surgery, 5 cardiac doctors and 4 nurses for the qualitative interview (Cohort 1) (**Table 1**), 500 patients for psychometric validation (Cohort 2), with median age of 56 years (IQR: 48-61) and 54 years (IQR: 44-61) (**Table 2**).

**Table 1.** Characteristics of participants in Cohort 1.

| Characteristics | Cohort 1 (n=40) |
| --- | --- |
| **Patients' level (N=31)** |  |
| Age, median (IQR), yrs | 56 (48-61) |
| <30 | 2 (6.5) |
| 30-40 | 2 (6.5) |
| 40-50 | 5 (16.1) |
| 50-60 | 12 (38.7) |
| ≥60 | 10 (32.3) |
| Female, No. (%) | 12 (38.7) |
| BMI, median (IQR), kg/m<sup>2</sup> | 25.3 (23.3-28.5) |
| Education, No. (%) |  |
| Less than high school | 15 (48.4) |
| High school | 8 (25.8) |
| University or above | 8 (25.8) |
| Type of surgery, No. (%)<sup>a</sup> |  |
| CABG | 15 (48.4) |
| Valve surgery |  |
| Mitral valve | 7 (22.6) |
| Tricuspid valve | 2 (6.5) |
| Aortic valve | 7 (22.6) |
| Aortic surgery | 7 (22.6) |
| Morrow | 2 (6.5) |
| Congenital | 2 (6.5) |
| Ablation | 2 (6.5) |
| Other <sup>b</sup> | 3 (9.7) |
| Official employment status, No. (%) |  |
| Unemployed | 7 (22.6) |
| Working or studying | 17 (54.8) |
| Retired | 7 (22.6) |
| Occupation type, No. (%) <sup>c</sup> |  |
| Physical (ISCO skill level = 1 or 2) | 11 (35.5) |
| Nonphysical (ISCO skill level = 3 or 4) | 6 (19.4) |
| Residential Area, No. (%) |  |
| Urban | 19 (61.3) |
| Rural | 12 (38.7) |
| Living alone status | 2 (6.5) |
| **Medical staff's level (N=9)** |  |
| Age, median (IQR), yrs | 38.0 (35.0-46.5) |
| Female, No. (%) | 5 (55.6) |
| Occupation, No. (%) |  |
| Doctor | 5 (55.6) |
| Nurse | 4 (44.4) |
| Status, No. (%) |  |
| Associate specialist | 5 (55.6) |
| Chief specialist | 4 (44.4) |
| Years of experience, median (IQR), yrs | 15.0 (10.0-23.0) |

Abbreviations: BMI, body mass index; CABG, coronary artery bypass grafting.
<sup>a</sup> Patients may have undergone multiple concurrent surgeries; therefore, the sum of type of surgery exceeds 100%. Percentages reflect the proportion of the cohort receiving each specific procedure type.
<sup>b</sup> The other 3 cases include 1 pulmonary artery thrombosis, 1 cardiac tumor, and 1 heart transplant.
<sup>c</sup> Classified according to International Labour Office

**Table 2.** Patient Characteristics in Cohort 2.

| **Characteristics** | **Cohort 2**<br>**(n=500)** |
| --- | --- |
| Age, median (IQR), yrs | 54 (44-61) |
| Female, No. (%) | 183 (36.6) |
| BMI, median (IQR), kg/m<sup>2</sup> | 24.7 (22.6-27.2) |
| Education, No. (%) |  |
| Less than high school | 218 (43.6) |
| High school | 112 (22.4) |
| University or above | 170 (34.0) |
| Comorbid conditions, No. (%) |  |
| Diabetes | 61 (12.2) |
| Hypertension | 175 (35.0) |
| Hyperlipidemia | 92 (18.4) |
| Smoking | 211 (42.2) |
| Alcohol consumption<br>(>1 time/week) | 197 (39.4) |
| LVEF, median (IQR), % | 65 (60-68) |
| LVEF < 45 | 10 (2.0) |
| LVEF ≥ 45 | 490 (98.0) |
| COPD | 7 (1.4) |
| Stroke | 11 (2.2) |
| Chronic renal failure | 5 (1.0) |

| Characteristics | Cohort 2<br>(n=500) |
| --- | --- |
| Peripheral arterial disease | 9 (1.8) |
| Prior cardiac surgery | 23 (4.6) |
| Type of surgery, No. (%)<sup>a</sup> |  |
| CABG | 113 (22.6) |
| Valve surgery |  |
| Mitral valve | 199 (39.8) |
| Tricuspid valve | 119 (23.8) |
| Aortic valve | 130 (26.0) |
| Aortic surgery | 71 (14.2) |
| Morrow | 73 (14.6) |
| Congenital | 91 (18.2) |
| Ablation | 24 (4.8) |
| Duration of surgery, median (IQR), min | 225 (187-276) |
| Hospital stay, median (IQR), d | 7 (6-7) |

Abbreviations: BMI, body mass index; LVEF, left ventricular ejection fractions; COPD, chronic obstructive pulmonary disease; CABG, coronary artery bypass grafting.
<sup>a</sup> Patients may have undergone multiple concurrent surgeries; therefore, the sum of type of surgery exceeds 100%. Percentages reflect the proportion of the cohort receiving each specific procedure type.

### Item generation

On the 23-item list in 9 domains (**Supplemental Table 1**) generated by the literature review (**Supplemental Figure 1**) and qualitative interview, a panel comprising 15 experts [7 clinicians, 7 nurses, and 1 researcher; 73.3% (11/15) with over 10 years in cardiac surgery] identified 17 items to generate the draft Fuwai-CRS. The instrument employed a 24-hour recall period and an 11-point scale ranging from 0 (not present) to 10 (as bad as you can imagine).

### Cognitive debriefing

21 patients in Cohort 2 underwent cognitive debriefing following the draft Fuwai-CRS assessment. Of these, 19 (90.5%) reported that the instrument was easy to complete, 19 (90.5%) deemed it easy to understand, and 20 (95.2%) stated that the 0-10 numeric scale was easy to understand. No other items were suggested in response to an open question posed to all patients **(Supplemental Table 2)**.

### Item finalization

We collected data from 500 patients in Cohort 2. Four items that exhibited a floor effect and one item exhibited a ceiling effect were recommended for elimination by the expert panel (**Table 3**). The results of hierarchical cluster analysis are presented in **Figure 1**. Based on the cluster analysis and clinical judgment, 3 items were eliminated. No items were eliminated in exploratory factor analysis. Finally, 9 items were left for further validation as the final Fuwai-CRS (**Table 4**). The flow chart of scale development is shown in **Figure 2**.

**Figure 1.**
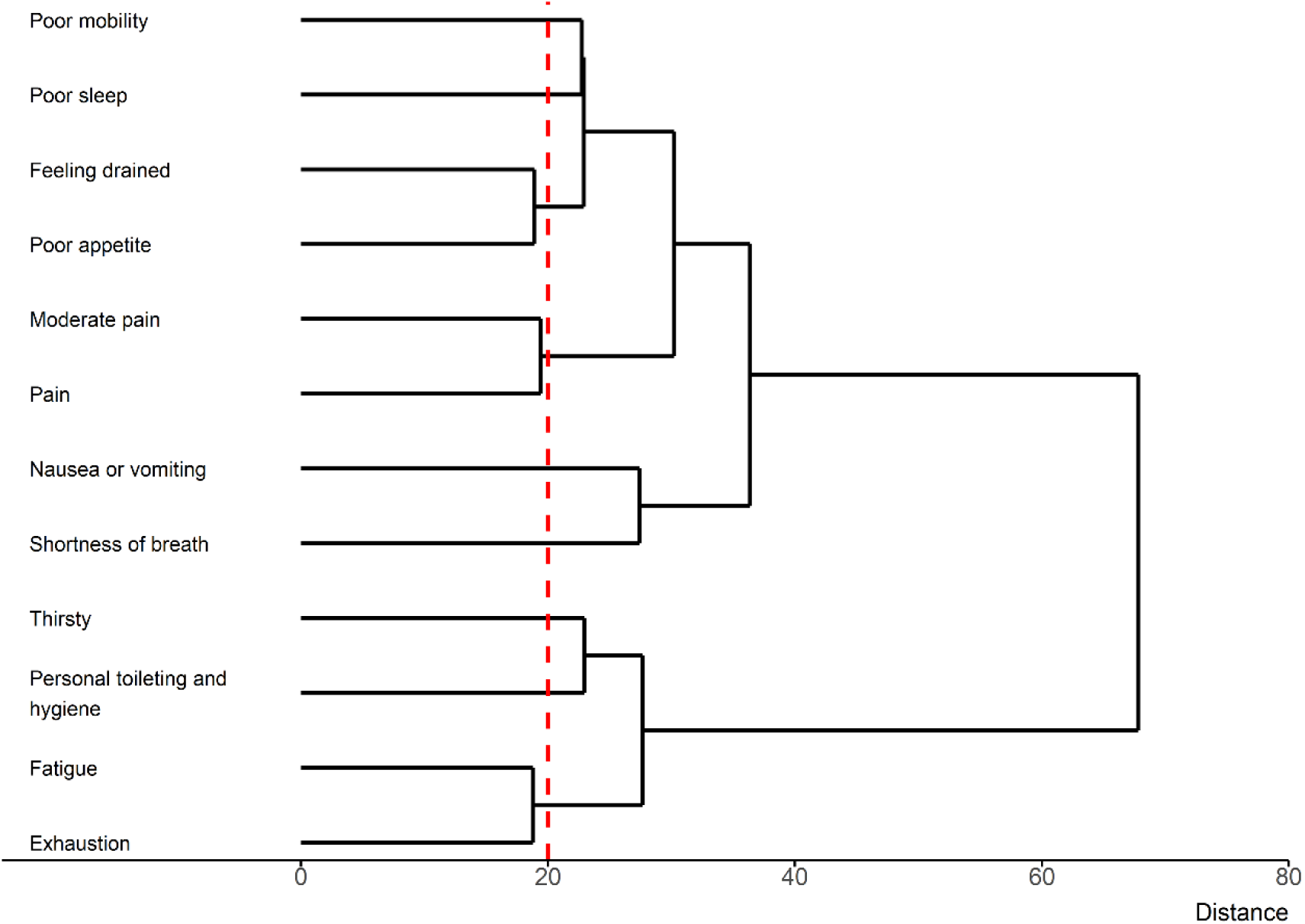
Relative distance among items. This dendrogram shows the results of a cluster analysis that was performed to examine the similarity of the items. Clusters were formed using the average linkage between groups, whereas the distances between items were calculated using squared Euclidian distances. The figure is read from left to right. Symptoms that join together earlier (toward the left side of the figure) are more similar than symptoms that join together later (toward the right side of the figure). The horizontal axis represents the distance between items, and similar items with a distance less than 20 will be simplified.

**Figure 2.**
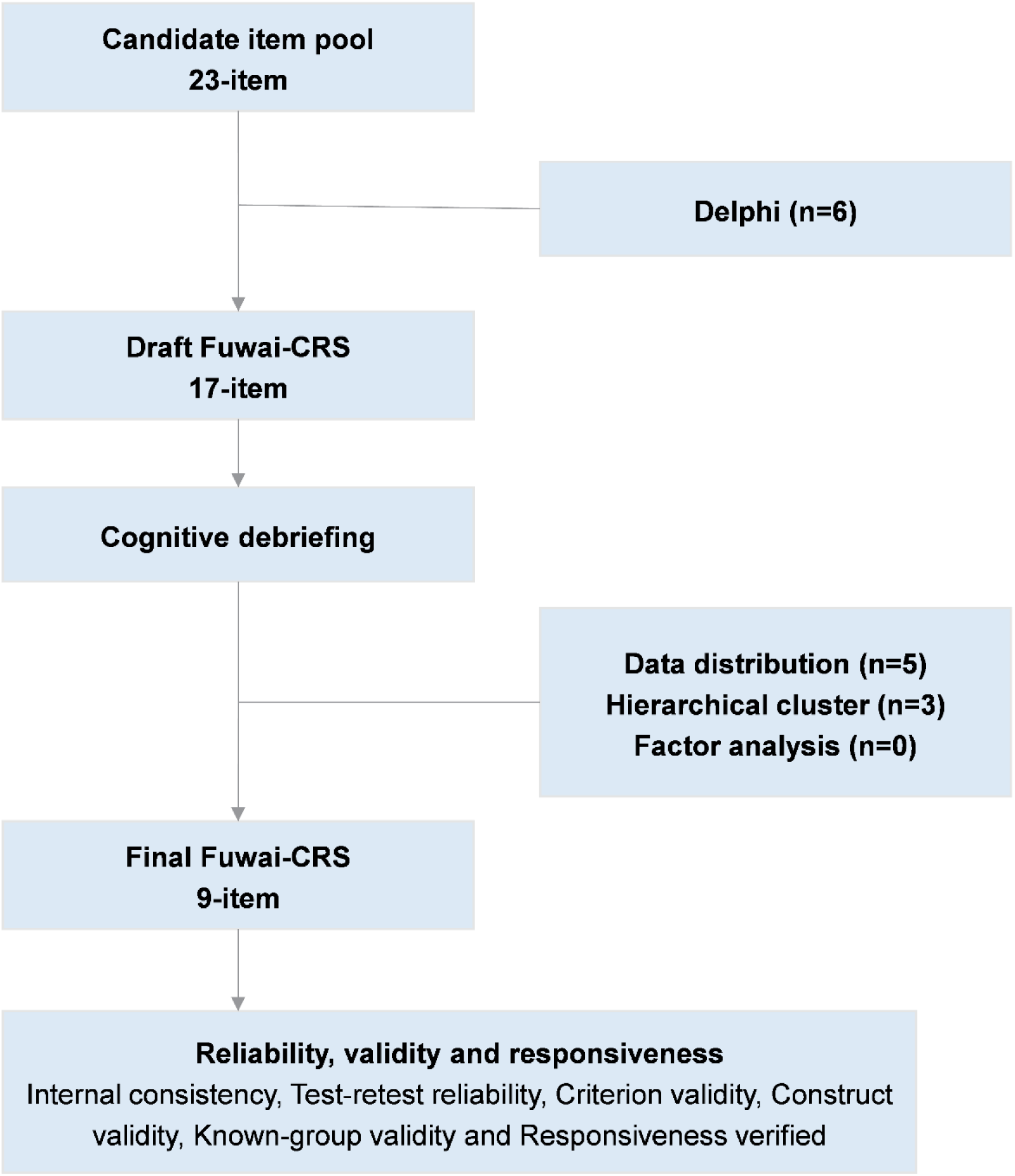
Development of Fuwai-CRS. Abbreviations: Fuwai-CRS, Fuwai Cardiac Recovery Scale.

**Table 3.** Item scores during hospitalization after surgery (Cohort 2)

| **Draft items** | **The average score of multiple time points <sup>a</sup> (n=500, Obs=1,734 <sup>b</sup>)** | **The most severe score of multiple time points <sup>a</sup> (Obs=500) <sup>c</sup>** | **Score=0 <sup>c, e</sup>** | **Score=10 <sup>d, e</sup>** |
| --- | --- | --- | --- | --- |
| Shortness of breath | 1.40 ± 1.69 | 2.51 ± 1.94 | 95 (19) | 0 |
| Poor appetite | 2.73 ± 2.64 | 4.51 ± 2.90 | 53 (10.6) | 0 |
| Feeling drained | 3.23 ± 2.58 | 5.09 ± 2.68 | 25 (5) | 0 |
| Poor sleep | 2.75 ± 2.51 | 4.65 ± 2.60 | 31 (6.2) | 0 |
| Unable to manage personal care independently | 6.86 ± 3.85 | 9.57 ± 1.73 | 4 (0.8) | 101 (20.2) |
| Unable to return to work or usual home activities <sup>f</sup> | 8.50 ± 2.69 | 9.83 ± 0.96 | 1 (0.2) | 202 (40.4) |
| Poor mobility | 4.66 ± 2.14 | 6.27 ± 1.77 | 0 (0) | 0 |
| Uncomfortable and out of control <sup>g</sup> | 1.08 ± 1.68 | 2.16 ± 2.14 | 160 (32) | 0 |
| Pain | 3.84 ± 2.79 | 5.60 ± 2.58 | 34 (6.8) | 0 |
| Moderate pain | 3.32 ± 2.79 | 5.39 ± 2.86 | 38 (7.6) | 1 (0.2) |
| Severe pain <sup>g</sup> | 0.83 ± 2.21 | 1.89 ± 3.13 | 336 (67.2) | 0 |
| Nausea or vomiting | 1.93 ± 2.67 | 3.55 ± 3.17 | 145 (29) | 2 (0.4) |
| Worried or anxious <sup>g</sup> | 1.68 ± 2.66 | 3.16 ± 3.35 | 217 (43.4) | 0 |
| Sad or depressed <sup>g</sup> | 1.36 ± 2.42 | 2.67 ± 3.17 | 249 (49.8) | 0 |
| Thirsty | 5.92 ± 3.47 | 8.50 ± 2.24 | 5 (1) | 29 (5.8) |
| Exhaustion | 5.00 ± 3.89 | 7.72 ± 3.10 | 41 (8.2) | 21 (4.2) |
| Fatigue | 5.57 ± 3.45 | 8.08 ± 2.53 | 20 (4) | 20 (4.0) |

Abbreviations: Obs, observations.
<sup>a</sup> Data is presented as means ± standard deviations. <sup>b</sup> Represents all completed assessments of patients during hospitalization. <sup>c</sup> Represents the most severe condition per patient during hospitalization. <sup>d</sup> Represents the best condition per patient during hospitalization. <sup>e</sup> Data is presented as n (%).
<sup>f</sup> Item with ceiling effect greater than 30 % is excluded. <sup>g</sup> Item with floor effect greater than 30 % is excluded.

**Table 4.**
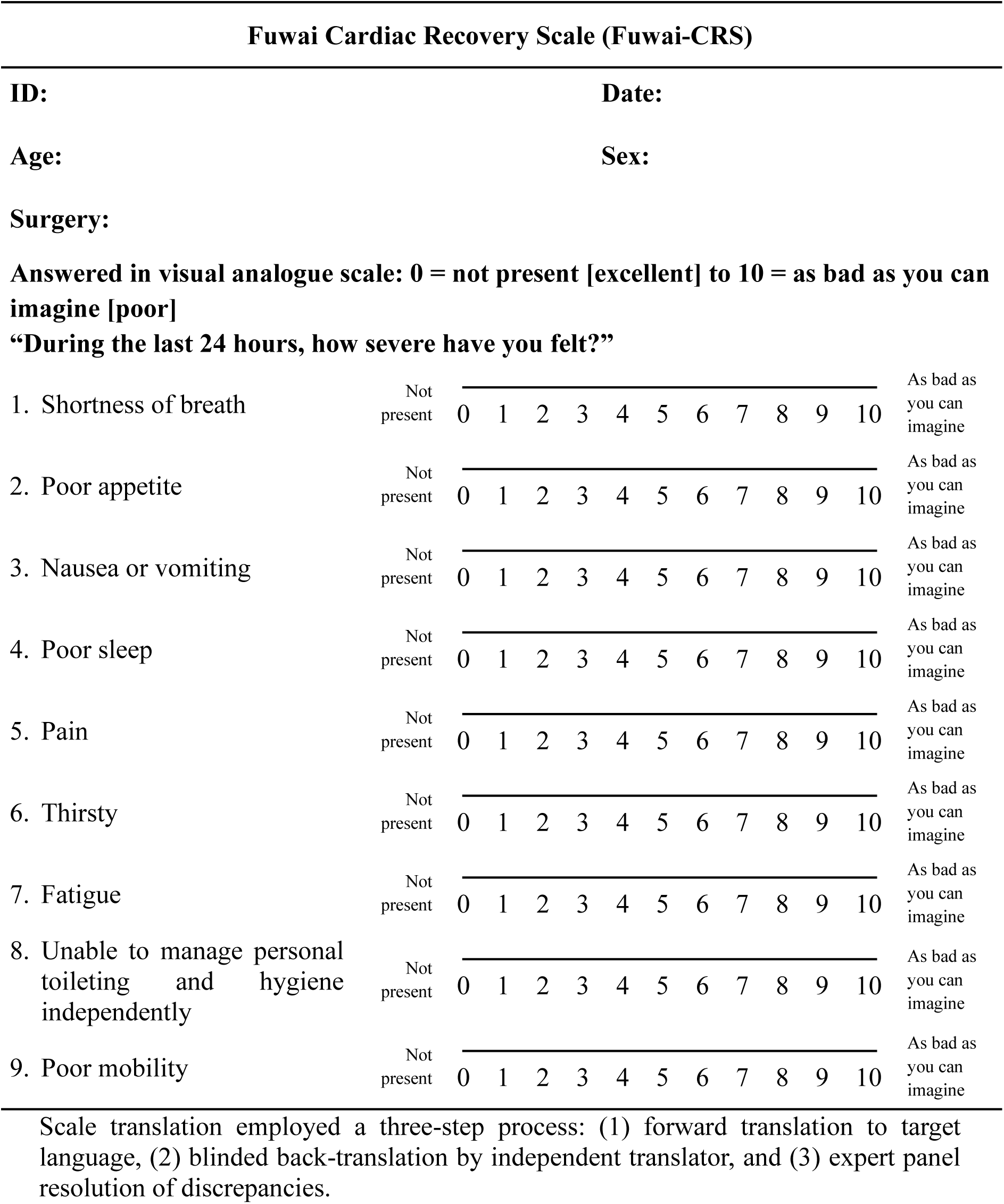
Final Fuwai-CRS.

### Reliability and validity

The final Fuwai-CRS showed good internal consistency (Cronbach’s α= 0.714) and test-retest reliability (ICC= 0.795). The Pearson’s correlation coefficients between Fuwai-CRS and QoR-15 was 0.875. Patients with preoperative LVEF values greater than or equal to 45 scored lower than those with LVEF values less than 45, and those with hospital stay after surgery greater than or equal to 7 days scored higher than those with hospital stay less than 7 days (**Supplemental Table 3**). The baseline and postoperative Fuwai-CRS scores were 8.3 ± 7.3 and 42.2 ± 13.0, respectively. This indicates excellent responsiveness, Cohen effect size of 4.66 and a standardized response mean of 2.53. Changes in perioperative scores and responsiveness are summarized in **Supplemental Table 4**.

## Discussion

Our study developed and validated a core-item scale (Fuwai-CRS) of postoperative recovery in patients undergoing cardiac surgery. The Fuwai-CRS was developed by a mixed-method study (Cohort 1 and Cohort 2) and was found psychometrically valid in the validation cohort (Cohort 2).

Systematic review demonstrated that evidence focusing on postoperative recovery of cardiac surgery was lacking and limited in quality.^27–29^ Significant heterogeneity were noted in tools (generic-surgery or cardiovascular-disease specific questionnaires) used to assess recovery.^29^ None of existed scales are developed or validated in representative cardiac surgery population (proportion of cardiac surgery in item generation cohort: QoR-40/QoR-15 0%, Post-operative quality recovery scale 5%).^30,31^ The present study enrolled a representative cardiac surgery population across different age, genders, types of surgeries, and socioeconomic status to develop a cardiac surgery specific PROM (Fuwai-CRS). The Fuwai-CRS was found to have good psychometrically validity, which filled the gap in this field.

Fuwai-CRS was designed to assess the core items in cardiac surgery recovery. Postoperative recovery status of cardiac surgery changes rapidly and is heterogeneous among different procedures. Since the frequent measurements are necessary, the item list should be minimally burdensome and reflect commonality concern in all types of cardiac surgery.^32^ Thus, we used standardized methodology in FDA guidance to include and exclude items for the clinical need (**Figure 2**).^16,17^ For example, previous studies demonstrate that emotional status represents an important domain for patients undergoing cardiac surgery, as the depression is reported to occur in 15-33% coronary artery bypass grafting (CABG) and 12.4% aortic valve replacement patients.^33,34^ Thus, we initially included four items addressing emotional domains during the item generation phase. However, the items of emotive domain were then removed by the floor effects (>30% of patients reporting zero from Day 1 to Day 4) of data distribution and Delphi panel’s exclusion criteria (**Table 3**). Our results are similar to the previous studies demonstrating that the postoperative emotive issues existed but not ubiquity, thus were removed.^33–36^ The final 9-item Fuwai-CRS can be completed in merely a minute with more than 80%-90% follow-up rate in our cohort (**Supplemental Table 5**). Future study is needed to assess how to properly append subset module of recovery items to the core Fuwai-CRS so that the fused subscale will more fit for specific patient condition.

Compared with previous generic scale (QoR-15), Fuwai-CRS newly included the domains of thirst and weakness, which demonstrated its cardiac surgery specific. Thirst after cardiac surgery is generated by the net negative fluid balance therapy to deal with the volume overload led by cardiopulmonary bypass or the heart dysfunction, which may not be concerned in non-cardiac surgery.^37^ Thirst measurement may be clinically useful as thirst-driven fluid management has been found effective in correcting abnormal fluid condition.^38,39^ Weakness initially included two items (fatigue and exhaustion), and subsequently only retained fatigue due to the hierarchical cluster analysis. Fatigue syndrome widely exists because of impaired cardiac function and inflammatory responses following cardiopulmonary bypass surgery.^40^ The presence of fatigue was ubiquity among cardiac surgery patients, which was associated with the recovery of physiologic function and physical activity.^41^ Standardized measurement and appropriate rehabilitation management may improve the fatigue syndrome.^40,41^

### Limitations

First, we recruited patients from hospitals in China, which may influence the study’s global generalizability. A broader, cross-cultural validation is needed in the future study. Second, the loss to follow-up questionnaires existed in 0%-26.71% patients, which happened more likely in the first two days (**Supplemental Table 5**). However, this would be acceptable compared with previous studies about perioperative PROMs assessment.^42,43^And the acceptable methodology design makes the present study enough to support our conclusions. Third, Fuwai-CRS was initially developed in Chinese version. Further studies are still warranted for multi-lingual scale development and validation.

## Conclusions

We developed and validated a core-item scale (Fuwai-CRS) to assess the postoperative recovery of cardiac surgery. The scale may serve as a promising tool for recovery assessment and management.

## Author Contributions

Concept and design: Zhe Zheng, Runchen Sun, Shen Lin, Chenfei Rao Methodology: Runchen Sun, Shen Lin, Shuang Hu, Qiuling Shi, Zhe Zheng Statistical analysis: Runchen Sun, Shen Lin, Zhongyu Jiao, Xiaoting Su Acquisition, analysis, or interpretation of data: Runchen Sun, Shen Lin, Chenfei Rao, Heng Zhang, Zhe Zheng

Project administration: Zhe Zheng, Yan Zhao, Sheng Liu, Wei Feng, Zhaoyun Cheng, Xiaoqi Wang, Chuzhi Zhou, Jue Wang, Yunpeng Ling, Zhenya Shen, Hai Tian Drafting of the manuscript: Runchen Sun, Shen Lin, Zhongyu Jiao, Zhe Zheng Critical review of the manuscript for important intellectual content: All authors.

## Ethics approval

This study was approved by Ethics Committee of Fuwai Hospital, CAMS&PUMC (reference number: 2022-1882).

## Data Availability Statement

The datasets used in this study are available from the corresponding author upon reasonable request.

