## Supplemental Materials for "Assessing the recovery after cardiac surgery: Development and validation of the Fuwai-CRS (Fuwai-Cardiac Recovery Scale)"

**Contents**


**Supplemental Methods**

**Supplemental Tables**

Supplemental Table 1. Candidate item pool

Supplemental Table 2. Cognitive debriefing details

Supplemental Table 3. Known-group validity: Scores by LVEF and Hospital stay (Cohort 2)

Supplemental Table 4. Change in the Fuwai-CRS score of patients before surgery (Preoperative baseline) and on the day after surgery (Postoperative)

Supplemental Table 5. Completion rate of scale collection

**Supplemental Figures**

Supplemental Figure 1. Literature review flow chart

### Supplemental Methods

#### Semi-structured interview

Standard in-depth semi-structured interviews were conducted with all participating patients, doctors, and nurses. The interviews included open-ended questions and subsequent probing questions, which were chosen based on previous literature review. Two experienced qualitative investigators with medical backgrounds conducted these interviews in a quiet and private space where the interviewees could share their views freely and confidentially without outside influence. One researcher conducted the interview, while the other took notes. All interviews were audiotaped and professionally transcribed.

In the qualitative analysis to identify the evaluation domains, qualitative data were analyzed and reported according to the Consolidated Criteria for Reporting Qualitative Research (COREQ) 32.[1] An inductive thematic analysis was applied to generate domains and items related to postoperative recovery of cardiac surgery. Two analysts coded each transcript independently and met to discuss the generation of domain/item and coding guidelines. All coding discrepancies were discussed and resolved by negotiated consensus. Analyses were performed with Nvivo V12.0 Plus qualitative software (QSR International Version 12, 2018).

For the qualitative study sample size, the concepts of information power and thematic saturation were used for purposive sampling. Interviews continued until there were no new concepts emerging from the data.

#### Literature review search terms

(("postoperative period"[MeSH Terms] OR (("postoperative"[All Fields] OR "post-operative"[All Fields]) AND "period"[All Fields]) OR "postoperative period"[All

Fields] OR "postoperative"[All Fields] OR "post surgical"[All Fields]) AND ("recovery"[All Fields]) AND ("cardiac surgical procedures"[MeSH Terms] OR "cardiac surgery"[All Fields] OR "cardiac surgical"[All Fields] OR "CABG"[All Fields] OR "coronary artery bypass"[All Fields] OR "valve replacement"[All Fields] OR "valve repair"[All Fields] OR "aortic"[All Fields] OR "morrow"[All Fields] OR "congenital"[All Fields] OR "ablation"[All Fields] OR "transplant"[All Fields])) AND English[Language]

The search criteria yielded 7042 studies. The inclusion and exclusion status of the studies were shown in Supplementary Figure 2. Finally, 239 articles were included for analyses.

**Supplemental Table 1. Candidate item pool**

| **Domain** | **Items** | **Sources of evidence** | |
|---|---|---|---|
| | | **Literature review (No. of studies)** | **Qualitative interview** |
| Physiology | 1. Shortness of breath | ✓ (89) | ✓ |
| | 2. Poor appetite | | |
| | 3. Feeling drained | | |
| | 4. Nausea or vomiting | | |
| Sleep | 5. Poor sleep | ✓ (62) | ✓ |
| Activity | 6. Unable to manage personal care independently | ✓ (102) | ✓ |
| | 7. Unable to return to work or usual home activities | | |
| | 8. Poor mobility | | |
| Social support | 9. Lacking support from hospital doctors and nurses | ✓ (68) | ✓ |
| | 10. Unable to communicate with family or friends | | |
| Emotion | 11. Uncomfortable and out of control | ✓ (139) | ✓ |
| | 12. Lacking a sense of general well-being | | |
| | 13. Worried or anxious | | |
| | 14. Sad or depressed | | |
| Cognition | 15. Unable to think clearly | ✓ (41) | ✓ |
| | 16. Unable to remember things | | |
| | 17. Unable to make decisions quickly | | |
| Pain | 18. Pain | ✓ (149) | ✓ |
| | 19. Moderate pain | | |
| | 20. Severe pain | | |
| Thirst | 21. Thirsty | × (0) | ✓ |
| Weakness | 22. Exhaustion | ✓ (14) | ✓ |
| | 23. Fatigue | | |

**Supplemental Table 2. Cognitive debriefing details**

| Questions | Evaluation criteria | Results (n=21) | |
|---|---|---|---|
| 1. How difficult do you think it is to complete the scale | Scored from 0 (no difficulty) to 10 (very difficult); scores >3 considered difficult, 0-3 considered easy | Easy to complete | 19 (90.5%) |
| | | Difficult to complete | 2 (9.5%) |
| 2. Do you have any items that you can't understand | "All understand" or "Part do not understand" | All understand | 19 (90.5%) |
| | | Part do not understand | 2 (9.5%) |
| 3. Do you understand the 0-10 rating? | "Yes" or "No" | Yes | 21 (100%) |
| | | No | 0 (0%) |
| 4. What do you think about the difficulty of using 0-10 score ? | Scored from 0 (no difficulty) to 10 (very difficult); scores >3 considered difficult, 0-3 considered easy | Easy to use | 20 (95.2%) |
| | | Difficult to use | 1 (4.8%) |
| 5. Other items not included in the scale | Open ended question | None | |

**Supplemental Table 3. Known-group validity: Scores by LVEF and Hospital stay (Cohort 2)**

| | **LVEF, % (LVEF ≥ 45 vs. LVEF < 45, 307 vs. 6)** | | | | **Hospital stay, d (≥ 7 vs. < 7, 88 vs. 234)** | | | |
|---|---|---|---|---|---|---|---|---|
| | Type | Score[a] | P value | Effect size | Type | Score[a] | P value | Effect size |
| Total score | LVEF ≥ 45 | 28.56 ± 13.14 | 0.03 | 0.92 | ≥ 7 | 33.98 ± 12.71 | <0.001 | 0.56 |
| | LVEF < 45 | 40.67 ± 17.11 | | | < 7 | 26.74 ± 12.95 | | |

Abbreviations: LVEF, left ventricular ejection fractions.

[a] Data is presented as mean ± standard deviations.

**Supplemental Table 4. Change in the Fuwai-CRS score of patients before surgery (Preoperative baseline) and on the day after surgery (Postoperative)**

| Fuwai-CRS Item | Preoperative [a] | Postoperative [a] | Mean Change (95%CI) | Cohen Effect Size [b] | Standardized Response Mean [c] |
|---|---|---|---|---|---|
| 1. Shortness of breath | 0.6 ± 1.0 | 1.4 ± 1.8 | 0.7 (0.5-0.9) | 0.69 | 0.38 |
| 2. Poor appetite | 0.9 ± 1.4 | 2.8 ± 3.2 | 1.9 (1.6-2.3) | 1.33 | 0.56 |
| 3. Nausea or vomiting | 0.2 ± 0.8 | 2.3 ± 2.7 | 2.1 (1.8-2.4) | 2.55 | 0.75 |
| 4. Poor sleep | 1.9 ± 1.9 | 2.6 ± 2.7 | 0.7 (0.4-1.0) | 0.37 | 0.23 |
| 5. Pain | 0.6 ± 1.5 | 3.7 ± 2.9 | 3.1 (2.8-3.5) | 2.06 | 1.00 |
| 6. Thirsty | 1.8 ± 2.5 | 8.0 ± 2.7 | 6.2 (5.8-6.5) | 2.46 | 1.74 |
| 7. Fatigue | 1.1 ± 2.3 | 5.9 ± 3.8 | 4.8 (4.4-5.2) | 2.06 | 1.14 |
| 8. Unable to manage personal toileting and hygiene independently | 0.2 ± 1.2 | 9.8 ± 1.0 | 9.6 (9.5-9.8) | 8.24 | 6.36 |
| 9. Poor mobility | 1.0 ± 1.5 | 5.7 ± 2.0 | 4.7 (4.4-4.9) | 3.14 | 1.96 |
| Total | 8.3 ± 7.3 | 42.2 ± 13.0 | 33.8 (32.5-35.2) | 4.66 | 2.53 |

Abbreviations: Fuwai-CRS, Fuwai Cardiac Recovery Scale.

[a] Data is presented as mean ± standard deviations. [b] Cohen Effect Size = mean change in score divided by the baseline (preoperative) standard deviation. [c] Standardized Response Mean = mean change in score divided by its standard deviation.

**Supplemental Table 5. Completion rate of scale collection**

| Time point | Patients who completed | Compliance Percentage (%)[a] |
|---|---|---|
| Baseline | 500 | 100 |
| D1 | 384 | 76.8 |
| D2 | 443 | 88.6 |
| D3 | 452 | 90.4 |
| D4 | 455 | 91.0 |
| W2 | 488 | 97.6 |
| W3 | 479 | 95.8 |
| M1 | 485 | 97.0 |
| M3 | 487 | 97.4 |

Abbreviations: Fuwai-CRS, Fuwai Cardiac Recovery Scale; D, day; W, week; M, month.

[a] The patient compliance is quantified by the ratio of the number of patients who successfully completed the Fuwai-CRS by the number of patients required to complete it at pre-determined time points.

**Supplemental Figure 1. Literature review flow chart**

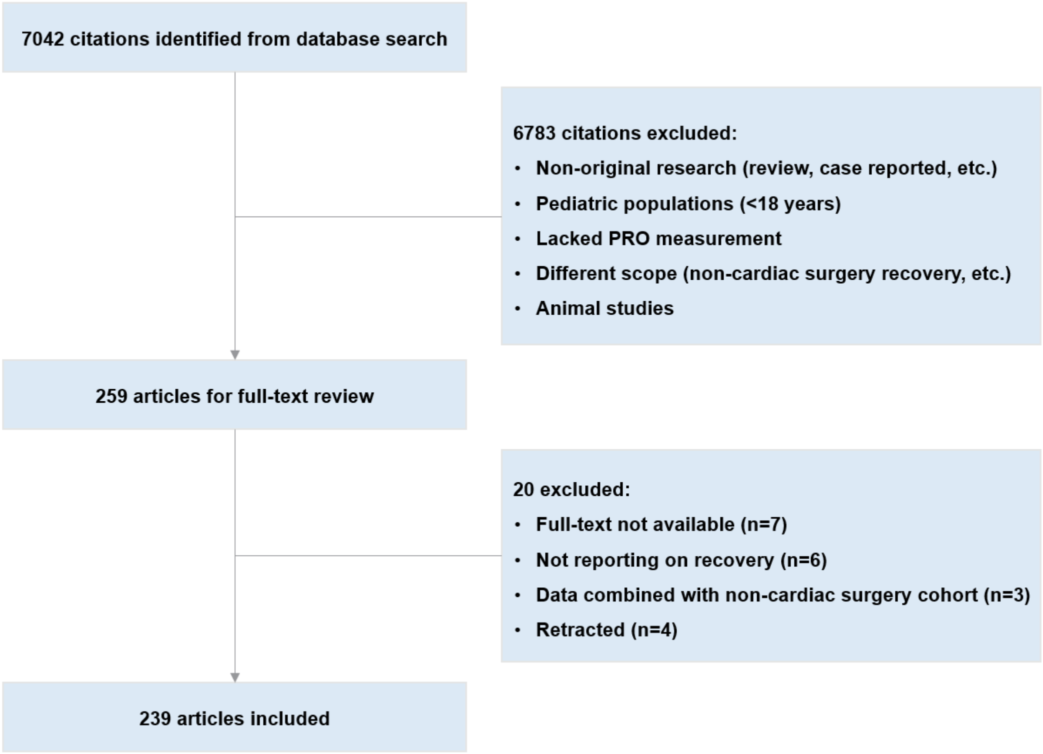


Abbreviations: PRO, patient-reported outcome.
